# Impact of lesion location and functional parameters on vision-related quality of life in geographic atrophy secondary to AMD

**DOI:** 10.1101/2023.09.22.23295946

**Authors:** Sandrine H. Künzel, Eliza Broadbent, Philipp T Möller, Moritz Lindner, Lukas Goerdt, Joanna Czauderna, Steffen Schmitz-Valckenberg, Frank G Holz, Maximilian Pfau, Monika Fleckenstein

## Abstract

**Background/Aims:** The primary objective was to determine how structural and functional parameters influence the vision-related quality of life (VRQoL) in patients with geographic atrophy (GA) secondary to age-related macular degeneration (AMD).

**Methods:** This prospective, non-interventional, natural-history ‘Directional Spread in Geographic-Atrophy’ study was conducted at the University Eye Hospital in Bonn, enrolling 82 patients with bilateral GA. Parameters such as GA location (assessed by the Early Treatment Diabetic Retinopathy Study grid), best-corrected visual acuity (BCVA), low-luminance visual acuity (LLVA), reading acuity, and speed were examined. The association between these parameters and VRQoL, as gauged using the National Eye Institute Visual Function Questionnaire 25 (NEI VFQ-25), was analyzed through least absolute shrinkage and selection operator with linear mixed-effects models.

**Results:** The average total GA area observed was 2.9 ± 1.2 mm^2^ (better eye) and 3.1 ± 1.3 mm^2^ (worse eye). The VRQoL scores for distance and near activities were most associated with the inner lower and inner left subfields of the better eye. For foveal-sparing patients, the LLVA of the better eye was the predominant determinate impacting all VRQoL scales.

**Conclusion:** GA location, specifically the inner lower and inner left subfields of the better eye, has a notable effect on VRQoL in GA patients. LLVA stands out as especially vital in foveal-sparing patients, underscoring the importance for clinicians to incorporate considerations of GA location and functional parameters into their risk-benefit assessments for emerging treatments.

**Key messenges:** *What is already known on this topic:* Despite recent therapeutic advancements, the relationship between GA markers and VRQoL remains unclear.

*What this study adds:* This research reveals the distinct value of specific structural and functional GA markers, notably the inner left and inner lower subfields, and LLVA, in relation to VRQoL in GA patients.

*How this study might affect research, practice, or policy:* Understanding these structural and functional markers will guide clinicians in personalized treatment decisions and encourage further research into more tailored therapeutic interventions.

## INTRODUCTION

Age-related macular degeneration (AMD) is a leading cause of visual impairment.^1,2^ In 2019, approximately 18.34 million US individuals aged 40 and older were living with early-stage AMD, and 1.49 million were living with late-stage AMD.^3^ Visual impairment caused by AMD compromises patient mobility and functioning. A retrospective study of 1901 patients with bilateral geographic atrophy (GA) secondary to AMD found that at initial diagnosis 71.1% did not meet the UK standard of visual acuity for driving, and 7.1% were legally blind. Within two years, an additional 66.7% became ineligible to drive, and 16% became legally blind.^4^ Further studies have reported that approximately one-fourth of cases of blindness in the US and UK are attributable to GA.^5,6^

GA refers to atrophic lesions that form on the retina due to pigment epithelium and photoreceptor degeneration.^7–9^ Lesions typically originate in the perifoveal macula and progress over time. Atrophy spreads more rapidly in the direction of the periphery of the retina than the fovea and may not involve the fovea until late in the disease.^10^ No preference has been found in the direction of spread from the center,^11^ and progression kinetics depend on the individual.^12,13^ This variability makes disease monitoring complex, as historically accepted markers of visual function in GA, such as BVCA and lesion size,^14–16^ may not directly correlate with patient impact.

For this reason, there has been a growing interest in using patient-reported outcome measures in GA. Vision-related quality of life (VRQoL) strives to capture patients’ experience of their disease, including its effects on daily functioning. Limited studies have examined the relationship between structural and functional markers and VRQoL in GA. Holm et al. found that BVCA and lesion size were associated with VRQoL.^17^ A study by our group reported a link between BVCA, GA size, and low luminance visual acuity (LLVA) of the better eye (BE) and VRQoL.^15^ In contrast, Ahluwalia et al. found no association between the total area of atrophy in the BE or worse eye (WE) and VRQoL. When examining the location of atrophy, involvement of the central 1-mm-diameter zone of the BE was associated with lower VRQoL.^18^ More research is needed to evaluate the relevance of structural and functional determinants of GA to VRQoL, especially in light of recent strides in therapeutics for AMD.

In 2023, pegcetacoplan and avacincaptad pegol were approved by the U.S. Food and Drug Administration (FDA) for the treatment of GA secondary to AMD. Both substances are repeatedly administered through intravitreal injections. They inhibit key steps in the complement pathway and have been shown to slow the enlargement of geographic atrophy lesions. ^19–21^

The yet uncertain risk-benefit ratio of these new drugs in clinical practice underscores the need for criteria to distinguish patients who may benefit from these novel therapeutic strategies from patients for whom these invasive, risk-associated interventions are highly unlikely to significantly affect their quality of life in a positive way.^22^

Our study investigates the relevance of a variety of GA determinants to VRQoL to aid clinicians in making this determination. To our knowledge, this paper is the first to analyze the relative contribution of structural and functional markers of GA to VRQoL in individuals with AMD.

## METHODS

### Patients

Patients were recruited at the Department of Ophthalmology at the University of Bonn, Germany, in the context of the Directional Spread in Geographic Atrophy study (DSGA, NCT02051998. Principal Investigator: Monika Fleckenstein). The DSGA study is a non-interventional, prospective, natural history study on GA secondary to AMD. This study adhered to the tenets of the Declaration of Helsinki and was approved by the institutional review board of the University of Bonn. Written informed consent was obtained, and participants received no stipend. To be included in this analysis, patients had to be at least 55 years of age at the baseline visit and have GA in both eyes.

Patients with exudative neovascular AMD or any other ocular disease that could affect the assessment of the retina in the study eye were excluded.

### Clinical Assessment

This study collected data on age, gender, medical history, BVCA, LLVA, reading acuity, foveal involvement, and NEI VFQ-25 scores. BCVA, LLVA (measured using a 2.0 log neutral density filter), and reading acuity were assessed using the Early Treatment Diabetic Retinopathy Study (ETDRS) and Radner Reading Charts, and confirmed to the base-10 logarithm of minimal angle of resolution (logMAR) scale.^23,24^ The NEI VFQ-25 includes a base questionnaire with 25 items that comprise a composite score and 12 subscales addressing various aspects of vision-related functioning. Scores range from 0 to 100, with higher scores indicating better function.^25,26^ In patients with neovascular AMD, a change of four to six points is considered clinically meaningful.^27^

### Imaging

After pupil dilatation using 0.5% tropicamide and 2.5% phenylephrine, patients underwent three types of imaging using a Spectralis HRA-OCT 2 (Heidelberg Engineering, Heidelberg, Germany): 30° × 30° fundus autofluorescence imaging (λ excitation, 488 nm; λ emission, 500–700 nm), 30° × 30° infrared reflectance imaging (λ, 815 nm), and 30° × 25° spectral-domain optical coherence tomography imaging (121 B-scans, ART 25).

### Image Grading

GA size was determined using the RegionFinder software for fundus autofluorescence (FAF) and infrared reflectance, as previously described, and the annotations were done in a semi-automatic manner.^28,29^ Two readers independently graded the images, and a third reader provided arbitration if the GA size differed by more than 0.3 mm^2^ between the first two readers. GA was classified as “foveal sparing” if the atrophic lesions did not include the center of the macula, and there was relative preservation of the outer retinal layers on OCT imaging subfoveally.

To assess the topographic location of GA, an ETDRS grid was placed onto the FAF images to assess the extent of GA within the subfields. The ETDRS grid consists of a central 1-mm-diameter zone, an inner 3-mm-diameter ring, and an outer 6-mm-diameter ring. The inner and outer rings are each divided into four quadrants (superior, inferior, nasal, temporal) to make 9 subfields, including the central zone.^30,31^

To simplify comparisons between eyes, subfields were labeled as they appear on retinal imaging (i.e., in retinal space based on the physician’s view). Subfields in the right eye temporal and left eye nasal quadrants are referred to as the left subfields (and correspond to the patient’s left visual field). Subfields in the right eye nasal and left eye temporal quadrants are referred to as the right subfields (and correspond to the patient’s right visual field). The labeling of the vertical quadrants of the ETDRS grid remained unchanged. The superior subfields correspond to the lower visual field and that the inferior subfields correspond to the upper visual field.

### Statistical Analyses

Statistical analysis of the data was carried out using R software and add-on packages lme4, glmnet, stepwise, and glmmLasso. GA size was square root transformed. Better and worse eyes were defined based on BCVA, and the Shapiro-Wilk test was used to assess variable normality. For normally distributed variables, the mean and standard deviation are presented. The median and interquartile range (IQR) are presented for nonnormally distributed variables. Univariable linear regression was applied to analyze the associations between the individual determinants and the dependent variable VRQoL.

Multicollinearity (two or more explanatory variables with high bivariate correlation) was evident, leading to instability in model coefficients and variable selection when using conventional multivariable least-squares regression. To address this issue, we applied least absolute shrinkage and selection operator (LASSO) regression with the VRQoL as the dependent variable for the cross-sectional multivariable analysis at baseline. LASSO regression is designed to handle multicollinearity and carries out variable selection by performing regularization and shrinking coefficient estimates toward zero. This enhances the prediction accuracy and interpretability by providing a parsimonious model (i.e., a model with few predictors).^32^

Nested cross-validation with patient-wise splits was applied to estimate the prediction accuracy of the model (outer leave-one-out cross-validation [LOOCV]) and to optimize the tuning parameter λ of the LASSO regression (nested inner LOOCV). For the longitudinal multivariable analysis, we applied LASSO regression (with nested cross-validation) for linear mixed-effects models with the VRQoL as the dependent variable considering the multicollinear and longitudinal data.^33^ Patients were considered as a random effect.

## RESULTS

A total of 164 eyes from 82 participants with GA were included in this analysis, with 43 females and 39 males. The mean age ± standard deviation (SD) was 77.2 ± 7.5 years of age at baseline. The average area of GA was 2.9 ± 1.2 mm^2^ for the BE and 3.1 ± 1.3 mm^2^ for the WE. The median (IQR) NEI-VFQ-25 composite score was 70 (25) (Table 1).

**Table 1.** Participant demographics, GA characteristics, and VRQoL performance. The cohort characteristics are shown. For normally distributed variables, the mean and standard deviation (SD) are shown. For non-normally distributed variables, the median and interquartile range (IQR) are given. Words per minute (wpm)

| DETERMINANT [UNIT] | MEAN ± SD |
| --- | --- |
|  | MEDIAN [IQR] |
| PARTICIPANTS | 82 (39 male, 43 female) |
| AGE [YEARS] | 77.2 ± 7.5 |
| BCVA BE [LOGMAR] | 0.30 [0.48] |
| BCVA WE [LOGMAR] | 0.84 [0.79] |
| GA AREA BE [MM] | 2.9 ± 1.2 |
| GA AREA WE [MM] | 3.1 ± 1.3 |
| FOVEAL SPARING AT LEAST ONE EYE [%] | 52.4% (43 eyes) |
| FOVEAL SPARING BOTH EYES [%] | 28% (23 eyes) |
| READING SPEED BE [WPM] | 92 [99] |
| READING SPEED WE [WPM] | 0.0 [95] |
| READING ACUITY BE [LOGRAD] | 0.63 [0.88] |
| READING ACUITY WE [LOGRAD] | 1.3 [0.69] |
| VRQOL COMPOSITE SCORE | 70 [25] |
| VRQOL NEAR ACTIVITIES SCORE | 50 [33] |
| VRQOL DISTANCE ACTIVITIES SCORE | 58 [33] |

Univariate and multivariate regressions were performed to understand ETDRS subfields most relevant for VRQoL. Figure 1 and Supplementary Figure S1 demonstrate the results of the univariate analysis in the left columns. Subfields of the BE tended to be more highly associated with VRQoL than those of the WE. Across composite and subscale scores, the most relevant subfields were the inner lower and inner left subfields of the BE. These subfields retained significance in the multivariate analysis, though the relative contribution of the inner left subfield to the model was greatest for the near activities subscale (Figure 1, Supplementary Figure S: right columns). For the composite score, the next most important subfields include the outer lower subfields of the BE and the inner lower subfield of the WE, which were also significant in the multivariate. For the near activities subscale, the next most relevant subfield was the inner left subfield of the WE. For the distance activities subscale, these included the inner lower subfield of the WE and inner right subfield of the WE.

**Figure 1.**
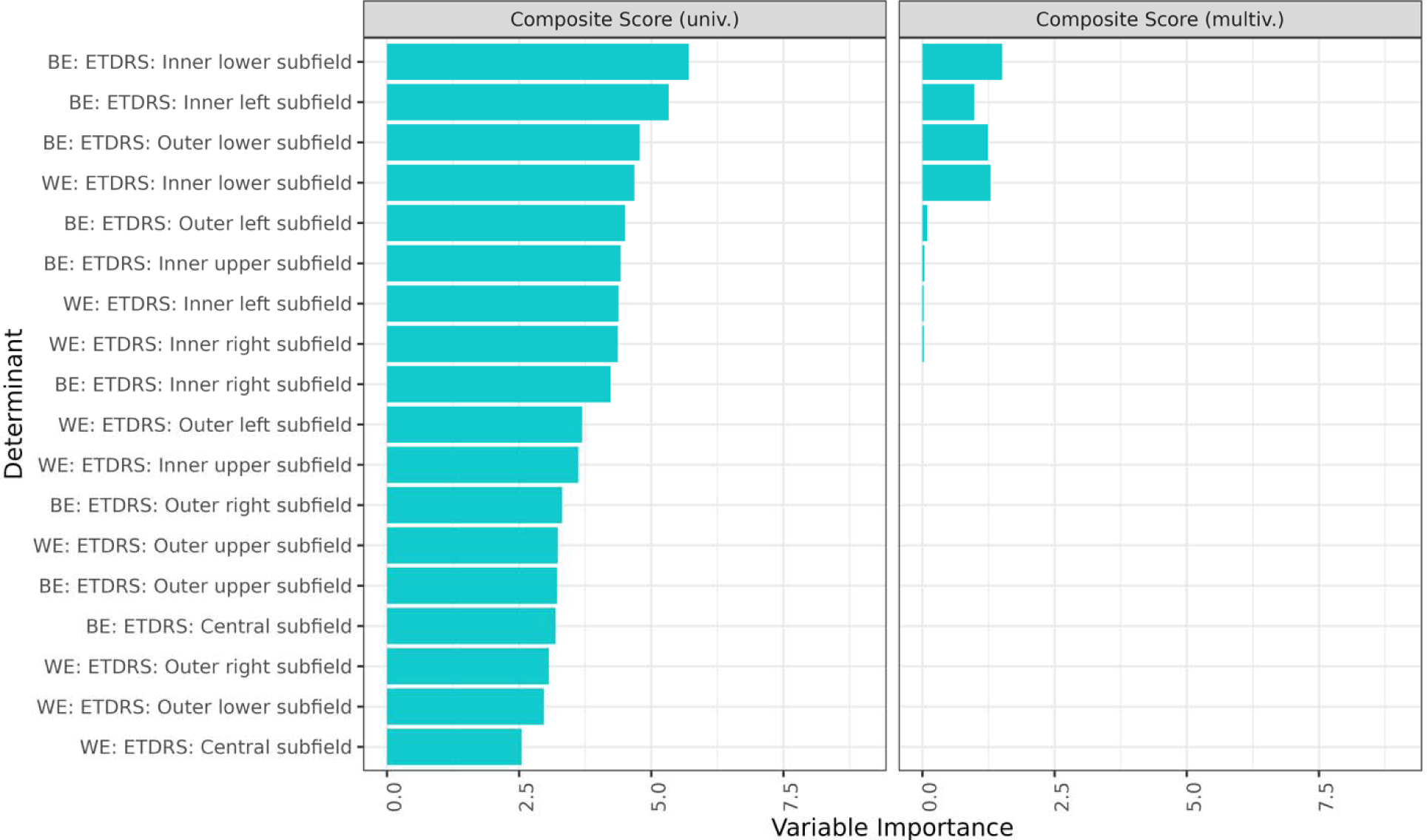
Uni- and multivariate regression of ETDRS subfields on VRQoL composite score. The variable importance was measured by the *t* statistic of the individual univariable linear mixed-effect models, and of the multivariable linear mixed-effect model with variables selected via least absolute shrinkage and selection operator regression for the univariate and multivariate analysis, respectively. BE, better eye; ETDRS, Early Treatment Diabetic Retinopathy Study; multiv., multivariate; univ., univerate; WE, worse eye

All structural determinants were then added. Results underscored the importance of the inner lower and inner left subfields of the BE to VRQoL, as these again emerged as most relevant in the univariate (Figure 2, Supplementary Figure S2, left column) and remained relevant in the multivariate (right column). GA left of fovea and total area of GA of the BE were also important to the composite and near activities subscales.

**Figure 2.**
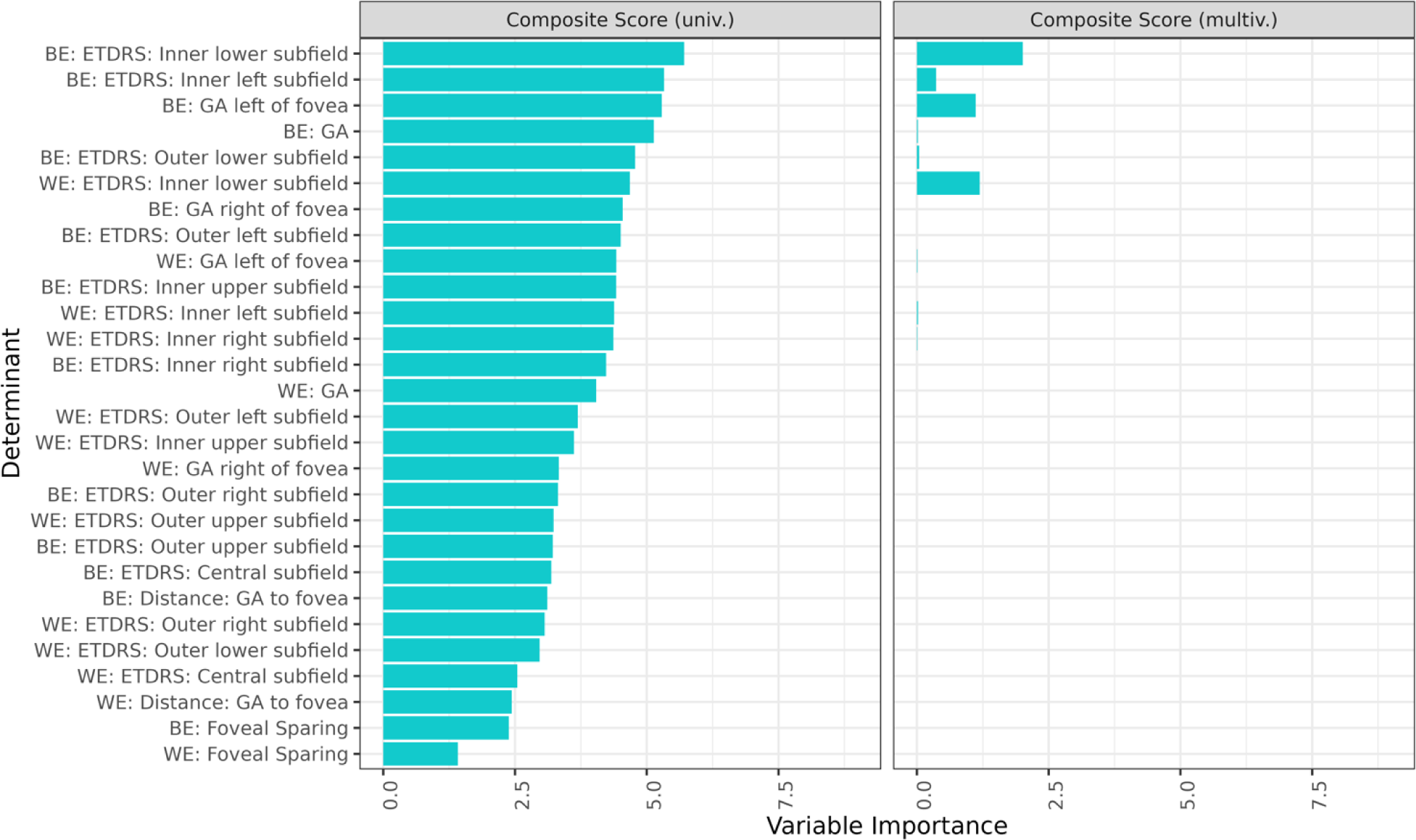
Uni- and multivariate regression of all structural determinants and ETDRS subfields with VRQoL. The variable importance was measured by the *t* statistic of the individual univariable linear mixed-effect models, and of the multivariable linear mixed-effect model with variables selected via least absolute shrinkage and selection operator regression for the univariate and multivariate analysis, respectively. BE, better eye; ETDRS, Early Treatment Diabetic Retinopathy Study; GA, geographic atrophy; WE, worse eye

Foveal-sparing GA of the better and WE were among the least relevant variables to composite and distance activities scores; however, foveal sparing demonstrated greater salience for near activities in the univariate and was the strongest contributor to the multivariate. For the distance activities scale, the subsequent most relevant determinants were the inner lower and inner right subfields of the WE.

Visual function variables were included to determine their comparative association with VRQoL with respect to structural factors. In the univariate regression, LLVA of the BE was the most salient factor for the composite scale, followed by the inner lower subfield, the inner left subfield, and reading speed, all of the BE (Figure 3, Supplementary Figure S3: left column). LLVA of the BE remained significant for the multivariate regression across all three scores, demonstrating its unique contribution to the model (right column). For the near activities scale, BCVA, reading acuity, and the inner left subfield of the BE were most relevant in the univariate. For the distance activities scale, the inner lower subfield, LLVA, inner left subfield, and reading speed of the BE were highly associated.

**Figure 3.**
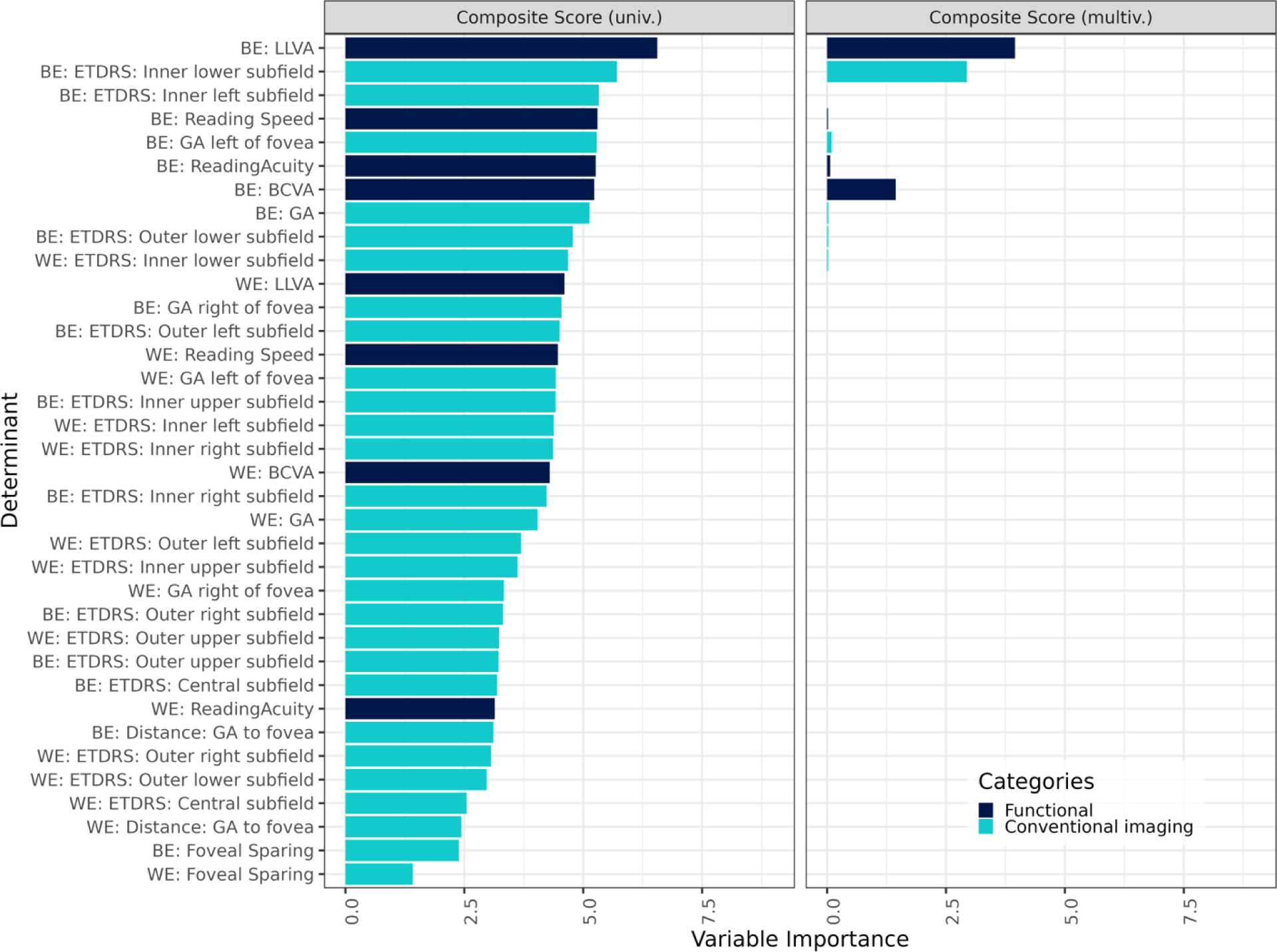
Uni- and multivariate regression of functional determinants, structural determinants, and ETDRS subfields with VRQoL. The variable importance was measured by the *t* statistic of the individual univariable linear mixed-effect models, and of the multivariable linear mixed-effect model with variables selected via least absolute shrinkage and selection operator regression for the univariate and multivariate analysis, respectively. BCVA, best-corrected visual acuity; BE, better eye; ETDRS, Early Treatment Diabetic Retinopathy Study; GA, geographic atrophy; LLVA, low luminance visual acuity; WE, worse eye

In a final approach, 86 eyes from 43 participants with foveal-sparing GA were analyzed separately. The univariate regression with only ETDRS subfields demonstrated the influence of the inner lower subfield of the BE to composite and distance activities scores, as well as the inner left subfield of the BE to near activities scores (Figure 4, Supplementary Figure S4). In the univariate regression with all variables, LLVA of the BE once again had the greatest relevance for composite scores in the univariate (Figure 5, Supplementary Figure 5). Additionally, it emerged as significant for both the near activities and distance activities subscales in the foveal-sparing cohort. The inner lower subfield remained important for the composite score, while reading acuity became relatively more important than reading speed for composite and distance activities scores within the univariate model.

**Figure 4:**
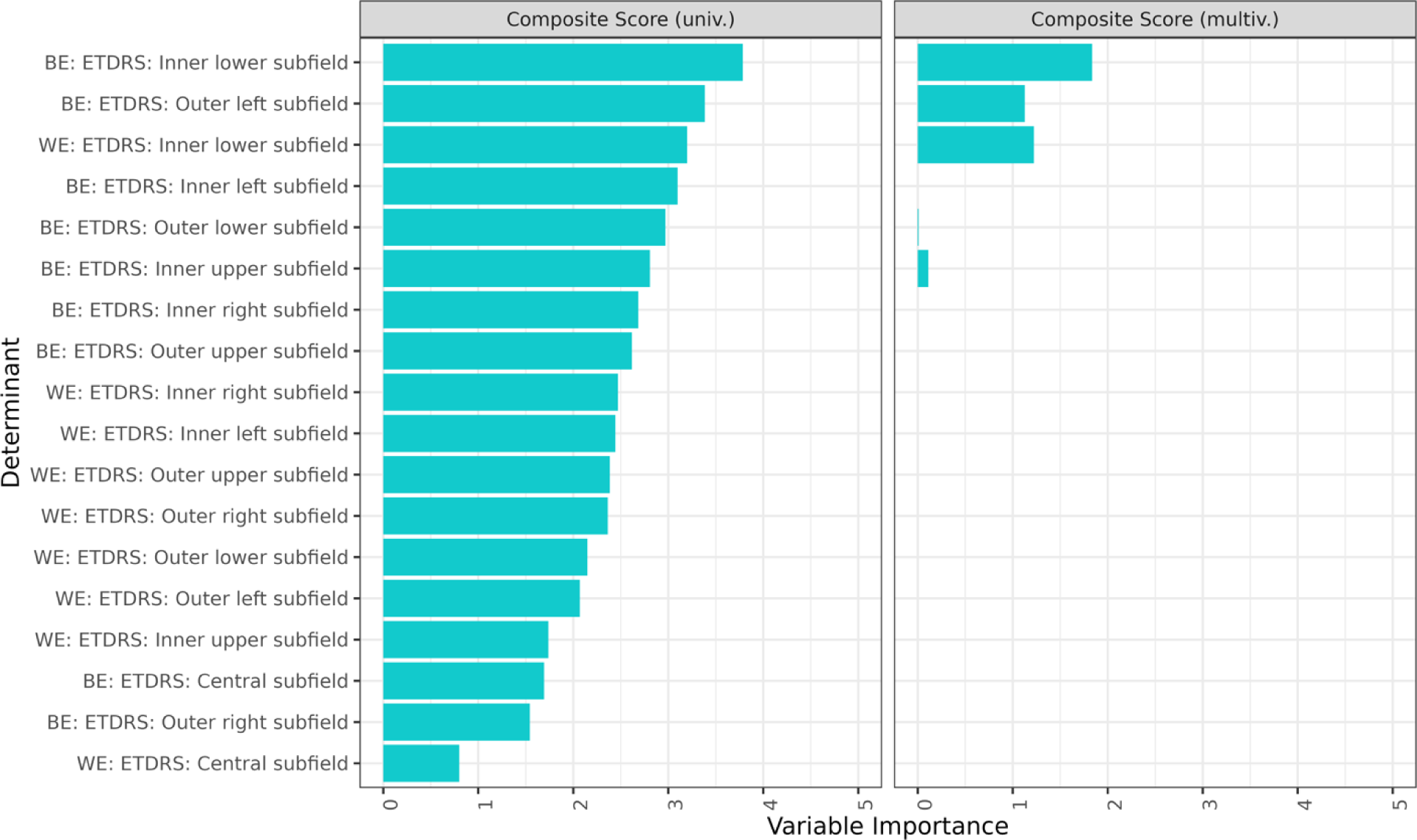
Uni- and multivariate regression of ETDRS subfields and VRQoL in foveal-sparing participants only. The variable importance was measured by the *t* statistic of the individual univariable linear mixed-effect models, and of the multivariable linear mixed-effect model with variables selected via least absolute shrinkage and selection operator regression for the univariate and multivariate analysis, respectively, each for the near activities (A) and distance activities (B). BCVA, best-corrected visual acuity; BE, better eye; ETDRS, Early Treatment Diabetic Retinopathy Study; GA, geographic atrophy; LLVA, low luminance visual acuity; WE, worse eye

**Figure 5.**
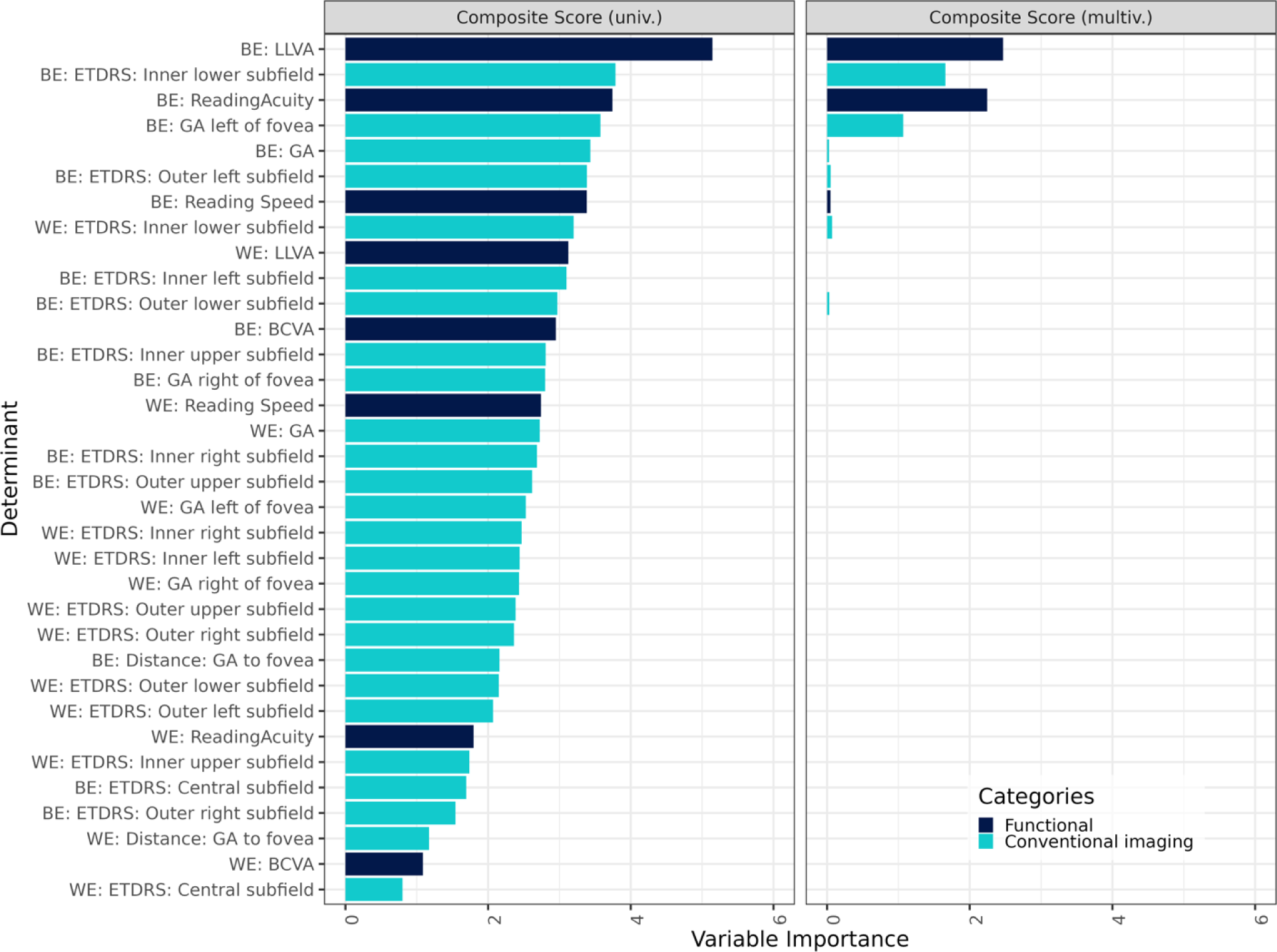
Uni- and multivariate regression of all variables and VRQoL in foveal-sparing participants only. The variable importance was measured by the *t* statistic of the individual univariable linear mixed-effect models, and of the multivariable linear mixed-effect model with variables selected via least absolute shrinkage and selection operator regression for the univariate and multivariate analysis, respectively. BCVA, best-corrected visual acuity; BE, better eye; ETDRS, Early Treatment Diabetic Retinopathy Study; GA, geographic atrophy; LLVA, low luminance visual acuity; WE, worse eye

## DISCUSSION

This study examined the relevance of GA determinants to VRQoL. Few studies have looked at ETDRS subfields in relation to quality of life outcomes,^18^ and to our knowledge, no prior study has shown the comparative salience of a wide range of structural and functional GA markers to different types of activities. Previously such findings had limited clinical applicability given the absence of FDA-approved treatment options.

In early 2023, the FDA approved pegcetacoplan for the treatment of GA. Pegcetacoplan is a complement protein C3 inhibitor that reduces GA progression when injecected intravitreally monthly or every-other-month ^32^ Another recently approved treatment, avacincaptad pegol (ACP) is a complement protein C5 inhibitor.^33^ Here, GA lesion growth showed reductions of up to 30.0%. Benefits of treatment were more pronounced across time points; a greater difference was seen between the treatment arm and the control arm at 18 months than at six months.^20^ Despite the morphological effects observed in the complement inhibitor studies, they have yet to show a significant impact on patient-reported outcome (PRO) assessments, possibly due to the small effect size or the unsuitability of the current PRO instruments, a contrast to their notable use in anti-VEGF studies in neovascular age-related macular degeneration (nAMD).

While the benefits of treatment appear clinically meaningful, they carry a burden for patients in terms of inconvenience, as injections appear to be most effective when administered frequently over a long period. As GA is often bilateral, patients may require injections in both eyes. Moreover treatment poses a dose-dependent risk of nAMD and ischemic optic neuropathy.^34^ There is ongoing uncertainty surrounding the risk-benefit profile of these new drugs in clinical practice.

Hence, it is imperative for practitioners and researchers to remain vigilant and informed about both the advantages and potential risks associated with these emerging treatments. These risks, which pose a potential threat to patients’ vision, underscore the critical importance of carefully choosing clinical markers to identify those who will truly benefit from the therapy.

Less-invasive treatments, such as dietary modification,^35^ may be more suitable for certain patient populations, making it critical to understand the likely course and implications of GA location in individuals. Previous research has shown that GA favors certain patterns of progression. GA tends to impact center and inner ETDRS zones, with most rapid enlargement in the inner zones. Increased area of GA positive correlates with outer zone involvement, with the superior outer subfield affected more frequently than other outer subfields.^11^ Localization of GA can be a powerful tool for clinicians in making treatment determinations, as specific subfields appear to be more critical for VRQoL.

Subfields of the BE tended to outperform their counterpart in the WE, consistent with prior studies.^15,18^ The inner lower and inner left subfields of the BE are especially important for VRQoL. When looking at structural determinants only, the inner lower subfield of the retina, corresponding to the superior visual field, was the greatest factor in the univariate and multivariate analysis for composite and distant activities scores. This subfield is implicated in activities such as driving (e.g., visualizing traffic signals), as found in the composite scale, and reading street signs or store names, as included in the distance activities subscale.

For the near activities subscale, the inner left subfield, referring to the left visual field, contributed most strongly to the univariate with structural determinants. This confirms Ahluwalia et al.’s finding that GA in the left visual field was significantly associated with lower VRQoL.^18^ As Western individuals typically read from left to right, the inner left subfield may play a role reading initiation. The left subfield has also been postulated to be important for moving to the next line while reading.^36^ Either of these functions could affect reading speed. Interestingly, while both the inner left subfield and reading speed were among the most relevant factors for the univariate regression with all variables, reading speed was no longer significant in the multivariate. Reading acuity and BVCA, by contrast, were significant to both. It is possible that GA in the inner left subfield subsumed the contribution of reading speed within the multivariate, suggesting the interrelatedness of these variables. This is supported by Keuster-Gruber et al.’s study of patients with hemaniopic field defects, in which horizontal training improved reading speed to a much greater extent in individuals with left field defects than those with right .^37^ More research is needed to examine this connection.

Of the functional markers, LLVA emerged as highly salient for VRQoL. While this was true for both samples analyzed, it was particularly evident for foveal-sparing GA. In the univariate analysis with all participants, LLVA was the most relevant variable for the composite score, while ranking slightly lower for distance activities and lower still for near activities. However, in the foveal-sparing analysis, LLVA was the most important variable for all scales in the univariate. Vision problems under low luminance are well-documented in elderly patients,^38–40^ and VRQoL related to low luminance activities is more likely to decline over time than for daytime activities.^40^ In patients with GA secondary to AMD, difficulty performing tasks under low luminance is seen at all stages of disease,^41^ and baseline deficit in LLVA is a strong predictor of future reductions in visual acuity.^42^ Sunness et al. identified adequate lighting as key for maximizing foveal vision in those with foveal-sparing scotomas.^43^ Previous studies have criticized the NEI VFQ-25 for focusing on photopic conditions to the neglect of low luminance.^40^ Even the significant role of LLVA in VRQoL illustrated in our results may be conservative due to inadequate representation of low-luminescence items. Future studies should incorporate scales targeted at low-luminescence activities^44,45^ to further investigate LLVA.

Strengths of this study were the variety of determinants used in the analysis, allowing for a holistic view of VRQoL, and the separate analysis of a foveal-sparing cohort. Limitations included the following: The sample size may limit generalizability and lack the statistical power to reveal weaker associations. The NEI NFQ-25, though validated for use in GA, relies on patients’ subjective self-report and may not perfectly capture VRQoL. Inclusion of an additional questionnaire targeted at low luminescence activities may have clarified the relationship between variables and VRQoL.

In conclusion, this study demonstrates the relative value of structural and functional GA markers to VRQoL. The inner left and inner lower subfields were most relevant for near activities and distance activities, respectively. Of the functional markers, LLVA was a notable contributor within the analysis. These findings can inform treatment decisions in regards to recently approved interventions for GA secondary to AMD.

## Data Availability

All data produced in the present study are available upon reasonable request to the authors

## Disclosures and Support

This work was supported by the German Research Foundation (DFG) under award numbers FL 658/4-1 and FL 658/4-2, the National Eye Institute of the National Institutes of Health under award numbers R01EY033365 and R01EY034965, the National Institutes of Health Core Grant (EY014800), an Unrestricted Grant from Research to Prevent Blindness, New York, NY, to the Department of Ophthalmology & Visual Sciences, University of Utah and the Bonfor O-137.0032 research grant of the university of Bonn to SK.

SK: Novartis Pharma (R), Chiesi GmbH (R), Apellis (R); EB: none; PM: none; ML: Bayer Healthcare (F); LG: BioEQ/Formycon (C), Novartis (R), BAYER Healthcare (R); JC: none; SSV: AlphaRET (C), Apellis (C, R), Bayer (F), Fomycon (C), Carl Zeiss MediTec (F), Galimedix (C), Heidelberg Engineering (F, R), Katairo (C), Kubota Vision (C), Novartis (C, F), Perceive Therapeutics (C), Pixium (C), Roche (C, F), SparingVision (C), STZ GRADE Reading Center (O); FH: Acucela (C,F), Alexion (C), Alzheon (C), Allergan (F), Apellis (C, F), Astellas (C), Bayer (C, F), Boehringer-Ingelheim (C), Bioeq/Formycon (F,C), CenterVue (F), Roche/Genentech (C,F), Geuder (C,F), Graybug (C), Gyroscope (C), Heidelberg Engineering (C,F), IvericBio (C, F), Janssen (C), Kanghong (C,F), LinBioscience (C), NightStarX (F), Novartis (C,F), Optos (F), Oxurion (C), Pixium Vision (C,F), Oxurion (C), Stealth BioTherapeutics (C), Zeiss (F,C), GRADE Reading Center (O); MP: Novartis (R), Janssen Pharmaceutica (C), Apellis (F,C); MF discloses to be Steffen Schmitz-Valckenberg’s spouse.

None of the above-mentioned entities had an influence on the design and conduct of the study; collection, management, analysis, and interpretation of the data; preparation, review, or approval of the manuscript; and decision to submit the manuscript for publication, respectively.

