## Supplemental Material for "Impact of lesion location and functional parameters on vision-related quality of life in geographic atrophy secondary to AMD"

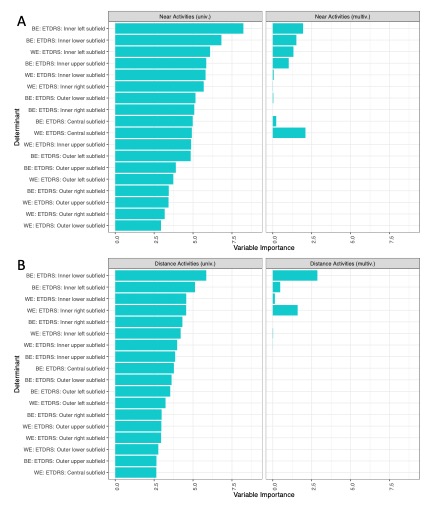

**Supplementary Figure S1. Uni- and multivariate regression of ETDRS subfields on VRQoL near and distance activities subscales.**

The variable importance was measured by the *t* statistic of the individual univariable linear mixed-effect models, and of the multivariable linear mixed-effect model with variables selected via least absolute shrinkage and selection operator regression for the univariate and multivariate analysis, respectively, Each for the near activities (A) and distance activities (B). BE, better eye; ETDRS, Early Treatment Diabetic Retinopathy Study; multiv., multivariate; univ., univerate; WE, worse eye

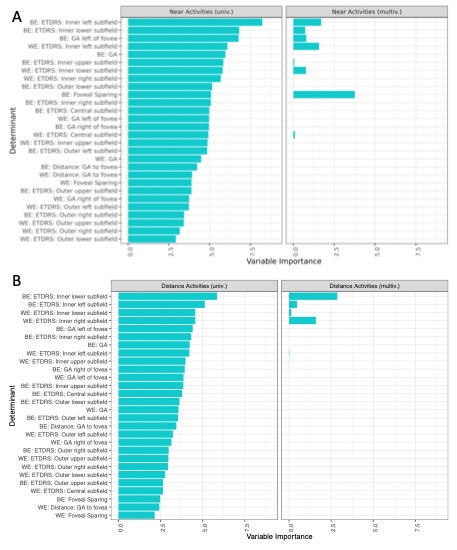

**Supplementary Figure S2.** **Uni- and multivariate regression of all structural determinants and ETDRS subfields with near and distance activities subscales.**

The variable importance was measured by the *t* statistic of the individual univariable linear mixed-effect models, and of the multivariable linear mixed-effect model with variables selected via least absolute shrinkage and selection operator regression for the univariate and multivariate analysis, respectively, each for the near activities (A) and distance activities (B). BE, better eye; ETDRS, Early Treatment Diabetic Retinopathy Study; GA, geographic atrophy; WE, worse eye

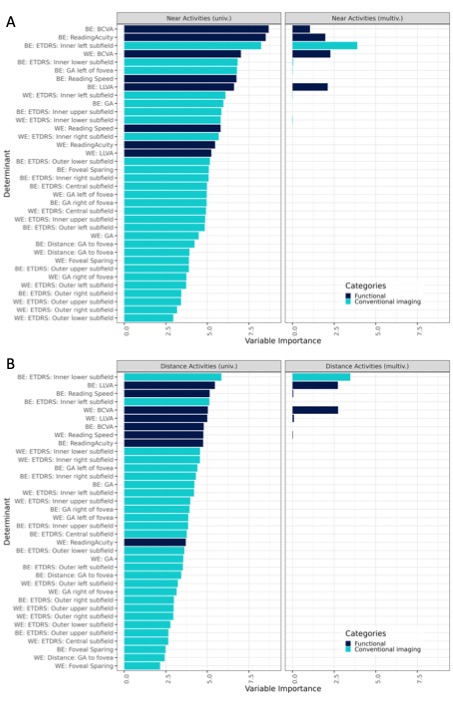

**Supplementary Figure S3.** **Uni- and multivariate regression of functional determinants, structural determinants, and ETDRS subfields with VRQoL for near and distance activities subscales.**

The variable importance was measured by the *t* statistic of the individual univariable linear mixed-effect models, and of the multivariable linear mixed-effect model with variables selected via least absolute shrinkage and selection operator regression for the univariate and multivariate analysis, respectively, each for the near activities (A) and distance activities (B). BCVA, best-corrected visual acuity; BE, better eye; ETDRS, Early Treatment Diabetic Retinopathy Study; GA, geographic atrophy; LLVA, low luminance visual acuity; WE, worse eye

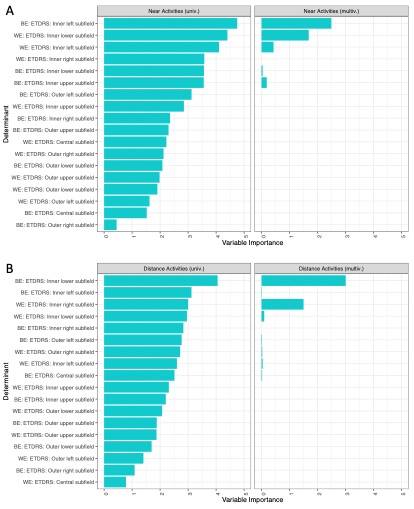

**Supplementary Figure S4: Uni- and multivariate regression of ETDRS subfields and VRQoL in foveal-sparing participants only for near and distance activities.**

The variable importance was measured by the *t* statistic of the individual univariable linear mixed-effect models, and of the multivariable linear mixed-effect model with variables selected via least absolute shrinkage and selection operator regression for the univariate and multivariate analysis, respectively, each for the near activities (A) and distance activities (B). BCVA, best-corrected visual acuity; BE, better eye; ETDRS, Early Treatment Diabetic Retinopathy Study; GA, geographic atrophy; LLVA, low luminance visual acuity; WE, worse eye

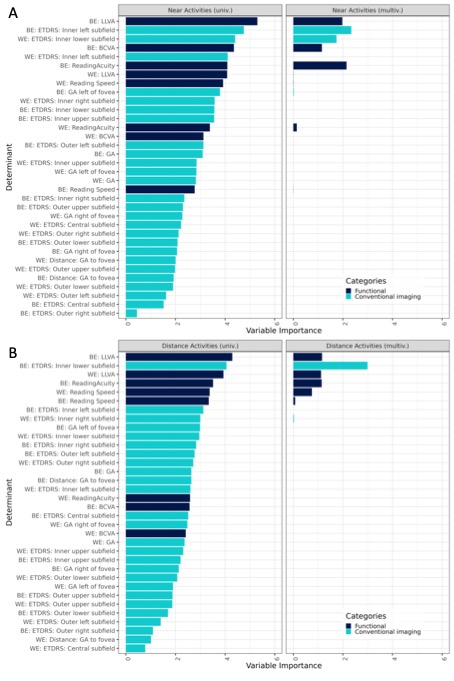

**Supplementary Figure S5.** **Uni- and multivariate regression of all variables and VRQoL in foveal-sparing participants only for Near and distance activities subscales.**

The variable importance was measured by the *t* statistic of the individual univariable linear mixed-effect models, and of the multivariable linear mixed-effect model with variables selected via least absolute shrinkage and selection operator regression for the univariate and multivariate analysis, respectively, each for the near activities (A) and distance activities (B). BCVA, best-corrected visual acuity; BE, better eye; ETDRS, Early Treatment Diabetic Retinopathy Study; GA, geographic atrophy; LLVA, low luminance visual acuity; WE, worse eye
